# Antibody repertoire associated with clinically diverse presentations of pediatric SARS-CoV-2 infection

**DOI:** 10.1101/2025.07.22.25331300

**Authors:** Natalie Bruiners, Rahul Ukey, Katherine C. Konvinse, Marlayna Harris, Muge Kalaycioglu, Jason H. Yang, Emily Yang, Usha Ganapathi, William Honnen, Tracy Andrews, Benjamin Richlin, Christian Suarez, Sunanda Gaur, Elizabeth Ricciardi, Uzma N. Hasan, William Cuddy, Aalok R. Singh, Hulya Bukulmez, David C Kaelber, Yukiko Kimura, Abraham Pinter, Stacey Napoli, Sandra Moroso-Fela, Lawrence C. Kleinman, Daniel B. Horton, Paul J. Utz, Maria Laura Gennaro

## Abstract

Pediatric SARS-CoV-2 infection results in clinical presentations ranging from asymptomatic/mild infection to severe pulmonary COVID-19, to Multisystem Inflammatory Syndrome in Children (MIS-C), characterized by hyperinflammation and multi-organ involvement. While various aspects of antibody responses to pediatric SARS-CoV-2 infection manifestations have been reported, parallel studies of antibody responses to viral and self-antigens are understudied. We tested whether clinical presentations of increasing severity corresponded to different antiviral antibody and autoantibody signatures. Using custom arrays, we found that, relative to uninfected subjects, all SARS-CoV-2 infection manifestations were associated with increased autoantibody production, suggesting pediatric SARS-CoV-2 infection as a risk factor for autoimmune complications. Subtle differences were seen in autoantibody patterns among infection groups, with some autoantibodies more associated with mild manifestations and others with severe ones. When we compared MIS-C and severe COVID-19 subjects, we found differences in IgG (mostly IgG1) abundance but not in Fc-mediated effector functions. Thus, MIS-C may be associated with abnormal antibody function, suggesting that this syndrome, and perhaps other post-acute sequelae of SARS-CoV-2 infection, may be associated with antibody dysfunction. Our study shows that the antibody repertoire varies with clinical presentation of SARS-CoV-2 in children and its analysis may help understand long COVID pathogenesis.

**Impact:** Our study shows that the antibody repertoire varies with the clinical presentation of SARS-CoV-2 in children, which has implications for understanding long COVID pathogenesis.

## Introduction

Infection of children with SARS-CoV-2 gives rise to a spectrum of clinical presentations. These range from asymptomatic or mild infection to severe COVID-19 and death. In addition, children infected with SARS-CoV-2 may present with a rare but severe post-acute complication referred to as Multisystem Inflammatory Syndrome in Children (MIS-C). MIS-C is characterized by hyperinflammation and multi-organ involvement ^1–5^, including cardiac dysfunction, abdominal pain, vomiting and diarrhea, and acute kidney injury ^6, 7^.

The antibody responses to SARS-CoV-2 infection in children have been investigated by many groups. Several studies have highlighted differences in durability, strength, and specificity of antibody responses between children and adults ^8–10^. These studies suggest that children exhibit responses that may contribute to their generally milder disease course and longer-lasting humoral immunity. Moreover, attention has been placed on autoantibody reactivity induced by SARS-CoV-2 infection. Recent research has identified autoantibody production in children following infection, particularly in those who develop MIS-C. The presence of autoantibodies in these children suggests immune dysregulation potentially linked to molecular mimicry or to other autoimmune mechanisms ^11–18^. However, parallel investigations in the same subjects of antiviral and autoantibody responses and their relationship to clinical severity of pediatric SARS-CoV-2 infection are lacking.

In the present study, we conducted analyses of antiviral and auto-antibodies in SARS-CoV-2-infected children presenting with clinical presentations ranging from asymptomatic and mild infection to severe COVID-19 requiring hospitalization, and to MIS-C. We investigated responses to structural and non-structural SARS-CoV-2 proteins, and we linked abundance and Fc-mediated effector functions to anti-spike antibodies. We also interrogated custom autoantibody arrays to identify profiles of antibody autoreactivity associated with occurrence and clinical severity of SARS-CoV-2 in children. Our results point to new hypotheses regarding antiviral antibody dysfunction and links between certain autoantibody markers and clinical severity.

## Materials and Methods

### Study Design and Population

COVID-19 Network of Networks Expanding Clinical and Translational approaches to Predict Severe Illness in Children (CONNECT to Predict SIck Children) is a multicenter prospective case-control study designed to predict children at greatest risk of severe consequences from SARS-CoV-2 infection. A total of 139 individuals who had confirmed infection and clinical signs of the disease were selected (**Table 1**). Cases were classified as asymptomatic/mild infection (*n* = 53), severe COVID-19 (*n* = 39), and MIS-C (*n* = 47), using the following criteria. Asymptomatic or mild SARS-CoV-2 infection cases included children and youth with a positive SARS-CoV-2 test who presented in the outpatient settings with no or mild symptoms (e.g., low-grade fever, sore throat, nasal congestion) not requiring inpatient care. Cases with severe COVID-19 were those requiring hospitalization for COVID-19-related pulmonary disease (e.g., pneumonia or acute respiratory distress syndrome [ARDS]). Children and youth with MIS-C were classified in accordance with the 2020 U.S. Centers for Disease Control criteria ^19^: (1) positivity for current or recent SARS-CoV-2 infection by RT-PCR, serology, or antigen test, or COVID-19 exposure within 4 weeks prior to onset of symptoms; (2) a combination of the following criteria: (a) fever; (b) laboratory evidence of inflammation (e.g., elevated CRP, D-dimer, IL-6); (c) evidence of clinically severe illness requiring hospitalization with dysfunction or disorders affecting at least 2 organs: cardiac, renal, respiratory, hematologic, gastrointestinal, dermatologic, or neurological; and (3) no alternative plausible diagnosis. Intravenous immunoglobulin treatment was administered to 33 MIS-C patients (70%). Information about COVID-19 vaccination status, and administration and timing of IVIG to MIS-C patients is provided in **Table 1**. In addition to samples from SARS-CoV-2-infected subjects, samples from 28 healthy pediatric controls (HC) were used. These included pre-pandemic samples (n =12) and samples obtained post-pandemic (n = 3) or at unspecified dates by a commercial source (Precision for Medicine, Gladstone, NJ, USA, and Discovery Life Sciences, Malden, MA, USA) (n = 13) that were serologically negative for SARS-CoV-2 Nucleocapsid.

**Table 1.** Demographic and clinical characteristics of subjects enrolled in the study.

|  | Total | 1. Mild/ Asymptomatic | 2. Severe COVID-19 | 3. MIS-C | P-value |  |
| --- | --- | --- | --- | --- | --- | --- |
| Covariate | N=139 | N=53 | N=39 | N=47 | 3 Groups | Group 2 vs 3 |
| Age |  |  |  |  | 0.34 | 0.32 |
| < 1 year old | 5 (3.60%) | 1 (1.89%) | 3 (7.69%) | 1 (2.13%) |  |  |
| 1-5 years old | 34 (24.46%) | 12 (22.64%) | 8 (20.51%) | 14 (29.79%) |  |  |
| 6-11 years old | 45 (32.37%) | 21 (39.62%) | 8 (20.51%) | 16 (34.04%) |  |  |
| 12-17 years old | 37 (26.62%) | 15 (28.30%) | 12 (30.77%) | 10 (21.28%) |  |  |
| 18-21 years old | 17 (12.23%) | 4 (7.55%) | 7 (17.95%) | 6 (12.77%) |  |  |
| Unknown | 1 (0.72%) | 0 (0.00%) | 1 (2.56%) | 0 (0.00%) |  |  |
| Sex |  |  |  |  | 0.047 | 0.65 |
| Female | 68 (48.92%) | 33 (62.26%) | 15 (38.46%) | 20 (42.55%) |  |  |
| Male | 70 (50.36%) | 20 (37.74%) | 23 (58.97%) | 27 (57.45%) |  |  |
| Unknown | 1 (0.72%) | 0 (0.00%) | 1 (2.56%) | 0 (0.00%) |  |  |
| Race/Ethnicity |  |  |  |  | 0.18 | 0.99 |
| Non-Hispanic White | 52 (37.41%) | 26 (49.06%) | 11 (28.21%) | 15 (31.91%) |  |  |
| Non-Hispanic Black | 24 (17.27%) | 6 (11.32%) | 8 (20.51%) | 10 (21.28%) |  |  |
| Non-Hispanic Asian | 11 (7.91%) | 7 (13.21%) | 2 (5.13%) | 2 (4.26%) |  |  |
| Hispanic | 40 (28.78%) | 12 (22.64%) | 13 (33.33%) | 15 (31.91%) |  |  |
| Missing | 12 (8.63%) | 2 (3.77%) | 5 (12.82%) | 5 (10.64%) |  |  |
| COVID vaccination status |  |  |  |  | 0.002 | 0.84 |
| Yes | 28 (20.14%) | 20 (37.74%) | 3 (7.69%) | 5 (10.64%) |  |  |
| No | 82 (58.99%) | 26 (49.06%) | 25 (64.10%) | 31 (65.96%) |  |  |
| Unknown | 29 (20.86%) | 7 (13.21%) | 11 (28.21%) | 11 (23.40%) |  |  |
| IVIG |  |  |  |  | <0.001 | <0.001 |
| Yes | 33 (23.74%) | 0 (0.00%) | 0 (0.00%) | 33 (70.21%) |  |  |
| No | 4 (2.88%) | 0 (0.00%) | 0 (0.00%) | 4 (8.51%) |  |  |
| Unknown | 102 (73.38%) | 53 (100.00%) | 39 (100.00%) | 10 (21.28%) |  |  |
| Days after hospitalization |  |  |  |  | <0.001 | 0.22 |
| 30 or Fewer: Incident | 55 (39.57%) | 6 (11.32%) | 25 (64.10%) | 24 (51.06%) |  |  |
| More than 30: Prevalent | 84 (60.43%) | 47 (88.68%) | 14 (35.90%) | 23 (48.94%) |  |  |

The p-values show the results of a comparison across all three diagnosis categories as well as a dichotomous comparison of severe COVID-19 and MIS-C. P-values were estimated using Pearson Chi-square Tests for sex, vaccination status, and days after the date of hospitalization; for age and race, Fisher Exact tests were used to account for small cell sizes.

### Study Activities and Data

Participants were enrolled between June 23, 2021, and November 24, 2022. Potential participants were identified through searches of electronic health records (EHRs) and recruited through direct contact in clinical settings or using flyers, phone calls, or emails. Following consent, parental permission, and/or assent as appropriate, participants with MIS-C or their parents or guardians completed a questionnaire. Clinical information (medical history, treatments, laboratory results, imaging, and other diagnostic tests) was obtained through EHR review. Peripheral blood was collected by phlebotomy and shipped to the study biorepository. Study data were collected and managed using REDCap electronic data capture tools hosted at the Rutgers Robert Wood Johnson Medical School ^20, 21^.

### Study approval

All study activities were approved by the Rutgers Institutional Review Board and other contributing healthcare systems through the reliant review process (Pro2020002961). All participants provided informed consent before engaging in study activities.

### Expression and purification of recombinant SARS-COV-2 proteins

Cloning and expression of the Receptor Binding Domain (RBD) of the S1 subunit of the SARS-CoV-2 Spike protein in the eukaryotic expression vector pcDNA3.4 (Addgene, Watertown, MA, USA) and 293-F eukaryotic cells was described previously ^22^. The entire SARS-CoV-2 Nucleocapsid (N) sequence (419 aa) was amplified and cloned in a pcDNA3.4 vector fused to a mouse leader sequence at the N terminal to enhance secretion and a Flag-H8 sequence at the C terminal to aid in purification. The resulting plasmid was transfected into 293F cells using the Expi293 Expression system (A14635, Thermo Fisher Scientific, Waltham, MA, USA), according to the manufacturer’s protocol. Supernatants were collected on day 4 post-transfection, and recombinant protein was purified by absorption to HisPur™ Cobalt Resin (89964, Thermo Fisher Scientific, Waltham, MA, USA) and elution with 200 mM imidazole. Purified protein was subsequently dialyzed against PBS at 4 °C. Absorbance (OD_280_) was determined by Nanodrop reading, and concentrations were calculated using the ExPASy Proteomics calculator.

### Enzyme-linked immunosorbent assay (ELISA)

Antibody binding was performed by ELISA utilizing recombinant SARS-CoV-2 Spike RBD and N proteins as solid-phase antigens and standard operating procedures, as described ^22^. Each sample was tested in duplicate. Plasma samples were used at a dilution of 1:80 for Ig isotype and 1:20 for IgG subclasses. Secondary antibodies included alkaline phosphatase-conjugated goat anti-human IgG antibodies (109-055-129, Jackson Immuno Research Laboratories, West Grove, PA, USA) used at 1:2,000 dilution for Ig Isotype detection and anti-human IgG subclasses (9052-04, 9070-04, 9210-04, 9200-04 and 2040-04, Southern Biotech, Birmingham, AL, USA) at a final concentration of 0.35 ug/mL.

### Cell lines

Reporter cell lines Jurkat-Lucia^TM^ NFAT-CD16 for Antibody-Dependent Cellular Cytotoxicity (ADCC) and Jurkat-Lucia^TM^ NFAT-CD32 for Antibody-Dependent Cellular Phagocytosis (ADCP) were obtained from InvivoGen (jktl-nfat-cd16, jkt-nfat-cd32, San Diego, CA, USA). Jurkat cells naturally express a functional nuclear factor of activated T cells (NFAT, a transcription factor that is involved in the early signaling events of ADCC and ADCP) ^23^. Moreover, these cell lines are engineered to express high-affinity CD16 and CD32. Engagement of these receptors by antibody Fc domain initiates a signaling cascade resulting in the expression of an NFAT-inducible Lucia luciferase reporter gene. Raji-Spike and Raji-Control cell lines were generously donated by Olivier Schwartz, Institute Pasteur, Paris, France ^24^. All cell lines were maintained in Roswell Park Memorial Institute (RPMI) 1640 medium (MT10104CV, Corning, NY, USA) supplemented with 10% heat-inactivated fetal bovine serum (97068-069, FBS; Seradigm, Radnor, PA, USA), 2mM L-glutamine (IC1680149, Corning Inc., Corning, NY, USA), and 1% penicillin/streptomycin (45000-653, Corning Inc., Corning, NY, USA), and maintained in a controlled environment at 37°C and 5% CO_2_ in a humidified atmospheric air environment. Blasticidin (10 µg/mL) and Zeocin (100 µg/mL) were employed for the maintenance of Jurkat-Lucia^TM^ cell lines.

For the ADCC and ACCP assays, 5×10^4^ Raji-Spike cells were co-cultured with 1×10^5^ Jurkat-CD16-NFAT (ADCC) or Jurkat-CD32-NFAT (ADCP) cells in the presence or absence of test sera, a dilution series of test or uninfected control serum. Luciferase was measured after 18 hrs (ADCC) and 6 hrs (ADCP) of incubation using a Synergy Neo2 BioTek microplate reader (Agilent, Santa Clara, CA, USA). ADCC and ADCP were expressed as fold-induction of luciferase activity relative to the ‘no serum’ control.

### Complement-Dependent Cytotoxicity (CDC)

Raji-Spike cells (5×10^4^) were seeded into a 96-well black, clear-bottom plate on the day before the experiment. On the day of the assay, Raji cells were incubated in the presence of 50% normal (NHS) or heat-inactivated (HIHS) human serum, with or without a dilution series of test or uninfected control serum. After 24h, cell lysis was determined using the CellTox Green Cytotoxicity Assay (Promega, Madison, WI, USA). CDC was expressed as fold induction of fluorescence activity compared to the ‘no serum’ control.

### Principal component analysis (PCA) and partial least squares discriminant analysis (PLS-PDA)

For analysis of data generated by ELISA and assays of Fc-mediated antibody functions, PCA and PLS-PDA were performed using custom scripts in MATLAB. Samples from each patient cohort were filtered to remove samples with antibody levels below detection, resulting in a total of n = 107 samples. Data were partitioned by SARS-CoV-2 infection state: prevalent vs. incident, MIS-C vs. severe COVID-19, vs. mild/asymptomatic SARS-CoV-2 infection. Samples with measurements above or below limits of detection were retained. Data were log10 transformed and standardized prior to PCA or PLS-DA analyses. Statistical tests for individual measurements were performed using the Welch’s t-test. *p* values of < 0.05 were considered significant.

### IgG antibody detection using a micro bead-based multiplex assay

A 119-plex protein array comprising a 73-plex autoantigen subarray and a 42-plex viral protein subarray was designed and fabricated as described, using a protocol adapted from previous work, with modifications ^1^. Briefly, antigens (**Supplemental Table 1**) were conjugated to uniquely bar-coded carboxylated magnetic beads (MagPlex-C, Luminex Corp, TX, USA). Post-conjugation, beads were validated using samples with known reactivity or monoclonal antibodies ^25^, aliquoted, snap-frozen in liquid nitrogen, and stored at –80°C until the day of the experiment, when they were thawed for use. Next, 45 μL of plasma, diluted 1:100 in PBS (46-013-CM, Corning) with 1% BSA (A3059, Sigma Aldrich, MO, USA), was added to a 384-well plate (Greiner BioOne) containing 5 μL of the bead array corresponding to ∼70 beads per antigen per well. Samples were incubated on a shaker at room temperature for 60 minutes, followed by an overnight incubation at 4°C. The following day, beads were washed with 0.05% PBS-Tween three times, using a plate washer (EL406, Biotek, VT, USA). Beads were then incubated with 50 μL of a 1:1,000 dilution of R-phycoerythrin–conjugated Fc-γ–specific goat anti-human IgG F(ab′)2 fragment (109-116-170, Jackson ImmunoResearch, PA, USA) for 30 minutes. After another round of washing with PBS-Tween, beads were resuspended in 50 μL of PBS-Tween for analysis. Analysis was performed using a FlexMap3D^TM^ instrument (Luminex Corp, TX, USA), and median fluorescence intensity (MFI) was measured. To validate the assay execution, some antigens were pre-qualified using either specific monoclonal antibodies or “Prototype” plasma samples from individuals who met classification criteria for known autoimmune diseases (e.g., Scl-70+ systemic sclerosis sera; Ro+ associated with lupus and Sjögren’s syndrome; Ribonucleoprotein+ associated with lupus and mixed connective tissue disease; and COVID-19 sera reactive with various viral antigens^25^. Samples were tested in duplicate, and MFI values obtained were normalized by subtracting the MFI values of bare beads.

### Analysis of micro bead-based multiplex assay data

For normalization, Python was used to average MFI values for “bare bead” IDs that were subtracted from average MFI values for antigen-conjugated bead IDs. The average MFI for each antigen was calculated using samples from healthy controls (HC). Antibodies were considered positive if MFI was >3000 units. In addition, we applied a stringent cutoff for “positivity” of >5 standard deviations (SD) above the average MFI for HC for the corresponding antigen. SD calculations and heatmaps were generated and visualized using R. For comparisons of MFI values among groups, a non-parametric Kruskal-Wallis test and Dunn-Bonferroni correction were performed. Analyses were performed and results displayed using GraphPad Prism. Adjusted *p* values of < 0.05 were considered significant.

## Results

### Anti-SARS-CoV-2 antibody profiles vary across the spectrum of clinical manifestations of SARS-CoV-2 infection

We measured IgG antibodies against viral antigens in three groups of SARS-CoV-2 infected participants (asymptomatic/mild infection, severe COVID-19, and MIS-C) and healthy (uninfected) controls utilizing a custom 42-plex protein array comprising SARS-CoV-2 structural and accessory proteins in addition to other common viral proteins (**Fig. 1A**, **Fig. S1A**, and **Table S1**). Statistically significant findings between healthy controls (HC) and SARS-CoV-2-infected patients were mainly limited to levels of IgG antibodies against immunodominant structural proteins [Spike receptor binding domain (RBD), Spike S1 subunit, Spike Ectodomain (ECD) S2 subunit, and Nucleocapsid], which were higher in all three infection groups relative to HC (**Fig. 1B-E**). Moreover, antibody levels for these structural proteins did not differ by infection severity status (**Fig. 1B-E**), except for antibodies against Spike S2 ECD that were significantly more abundant in the mild SARS-CoV-2 infection than in severe COVID-19 group (**Fig. 1E**).

**Figure 1.**
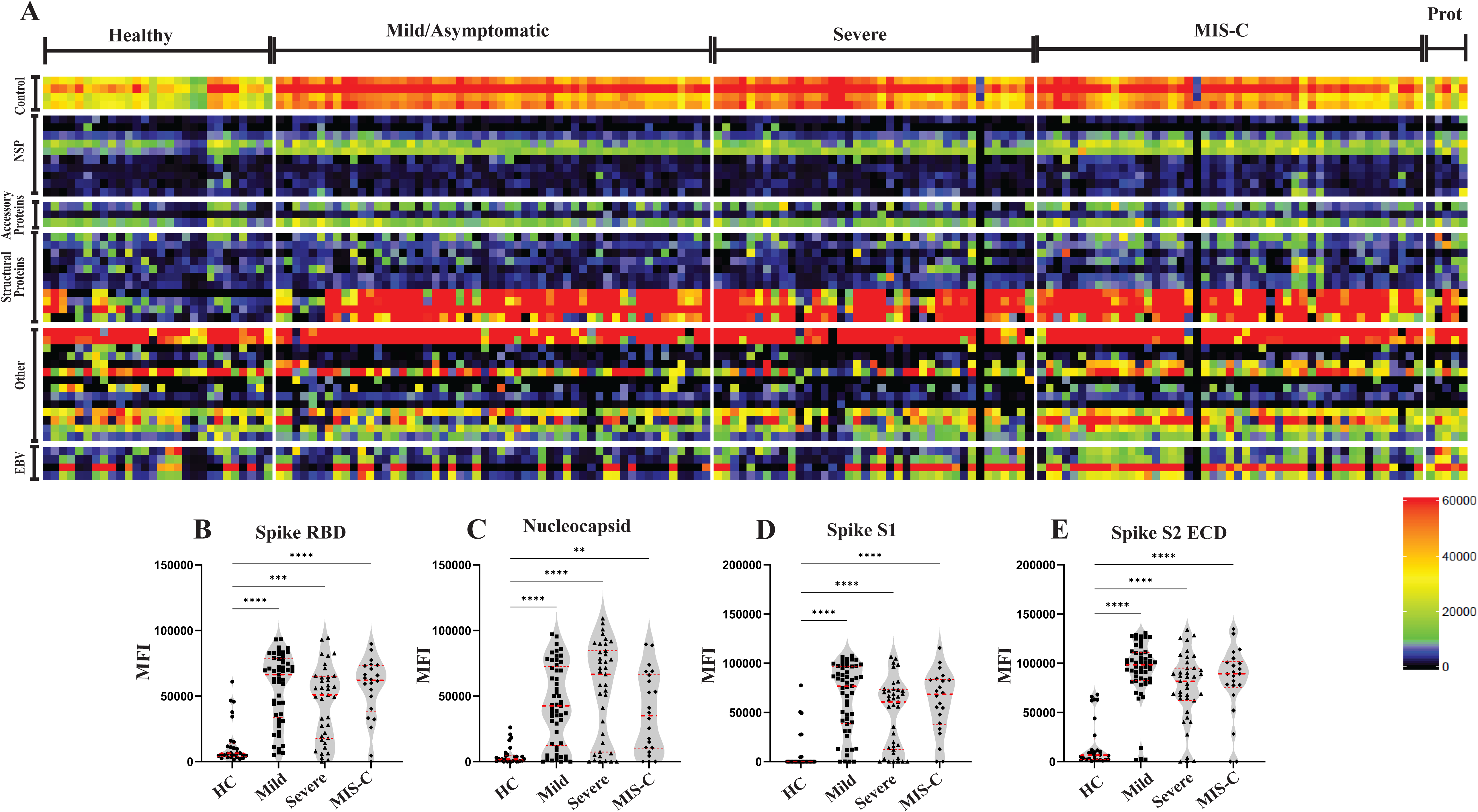
Plasma IgG antibodies against SARS-CoV-2 structural proteins but not nonstructural proteins or proteins from other viruses are elevated in patients with varying SARS-CoV-2 infection severities compared to healthy controls (HC). (A) Heatmap representing serum IgG measured using a 42-plex protein array. Healthy controls (HC) (n = 28), and patients who were diagnosed with mild/asymptomatic SARS-CoV-2 infection (n = 53), severe COVID-19 (n = 39), and MIS-C (n = 47), and prototypes (positive controls) (n = 5) are shown on the x-axis. Antigens are grouped on the y-axis by SARS-CoV-2 subgroups (Non-structural and structural proteins), and other viral antigens, including EBV. Antigen names are provided in **Table S1**. Colors correspond to MFI gradient values. (B-E) Violin plots comparing MFI data among HC, mild SARS-CoV-2 infection, severe COVID-19, and MIS-C patients who were at least 100 days post-IVIG treatment (n = 18). The dotted middle line is the median, and the upper and lower dotted lines correspond to the first and third quartiles. MFI values are represented on the y-axis. MFI values for individual participants are displayed as dots for HC, squares for mild infection, triangles for severe COVID-19, and diamonds for MIS-C. Violin plots show IgG measurements of key SARS-CoV-2 structural proteins: SARS2 Spike RBD (B), SARS2 Nucleocapsid (C), SARS2 Spike S1 (D), SARS2 Spike S2 ECD (E). Dunn-Bonferroni statistical analyses and corrections were performed to determine statistical significance between groups, with asterisks corresponding to the adjusted p values; **** p <0.0001, *** p <0.001, ** p <0.01, * p <0.05. Non-significant comparisons (p > 0.05) are not shown. Source data are provided as a Source Data file.

In addition to the structural proteins, antibodies against two SARS-CoV-2 associated antigens [nonstructural protein (NSP15) and open reading frame (ORF8)] were also higher in the three infection groups relative to HC (**Fig. S1B**). NSP15 is involved in facilitating RNA processing ^26^, and ORF8 has roles in viral replication and immune evasion^27^.Moreover, antibodies against two additional nonstructural proteins (NSP2, NSP3) were increased only in mild/asymptomatic infection (NSP2) and in MIS-C (NSP3), relative to HC (**Fig. S1C**). Furthermore, we detected no differences between infected and HC groups in IgG antibody levels to accessory proteins such as ORF8, ORF6, and ORF7a, which are involved in inhibition of type 1 interferons ^28^ (**Fig. S1D**). Interestingly, we also observed that IgG antibodies against Bcl-2 interacting protein 3 (BNIP3), a protein involved in cell death, and SARS-CoV-2-PLpro (papain-like protease), an enzyme required for viral polyprotein maturation, were higher in HCs relative to all infected groups (**Fig. S1E**). Together, these findings highlight a complex and heterogeneous antibody response to SARS-CoV-2 in children, marked by conserved reactivity to structural proteins across clinical phenotypes, alongside selective modulation of responses to nonstructural and accessory proteins that may reflect differential immune activation or regulation associated with disease severity.

### Abundance, but not Fc-mediated effector functions, of anti-Spike IgG antibodies differ between incident severe COVID-19 and MIS-C

For further analysis of the anti-SARS-CoV-2 antibody response, we used ELISA to assess the abundance of total IgG to Spike RBD and Nucleocapsid (N), and IgA, IgM, and IgG subclasses against Spike RBD. Moreover, to explore the biological functions of anti-Spike antibodies in children, we examined their ability to elicit Fc-mediated effector functions such as antibody-dependent cellular cytotoxicity (ADCC), antibody-dependent cellular phagocytosis (ADCP) and complement-dependent cytotoxicity (CDC). To account for time elapsed between manifestation of disease and blood draw obtained for the study, we subdivided subjects with severe COVID-19 and MIS-C in incident (I, when blood was collected within 30 days from hospital admission) and prevalent (P, when blood was collected >30 days from hospital admission) (**Table 1**). Using principal component analysis (PCA), we found no differences in IgG abundance and function among mild/asymptomatic infection, and prevalent severe COVID-19 and MIS-C (**Fig. 2A**). It was noted however that incident MIS-C patients appeared to segregate from incident severe COVID-19 (**Fig. 2A**). We next performed partial least squares discriminant analysis (PLS-DA) to create a model that classifies these two groups based on all antibody measurements. We found that indeed the model distinguishes between severe COVID-19 and MIS-C (**Fig. 2B**, left panel). Analysis of the distinguishing variables in the model showed that anti-RBD IgG antibody levels were associated with MIS-C while anti-N antibodies tended to be associated with severe COVID-19 (**Fig. 2B**, right panel), consistent with the results obtained with the multiplex array platform (**Fig. 1B**). In contrast, the Fc-mediated effector functions showed no predictive value in differentiating between the two groups.

**Figure 2.**
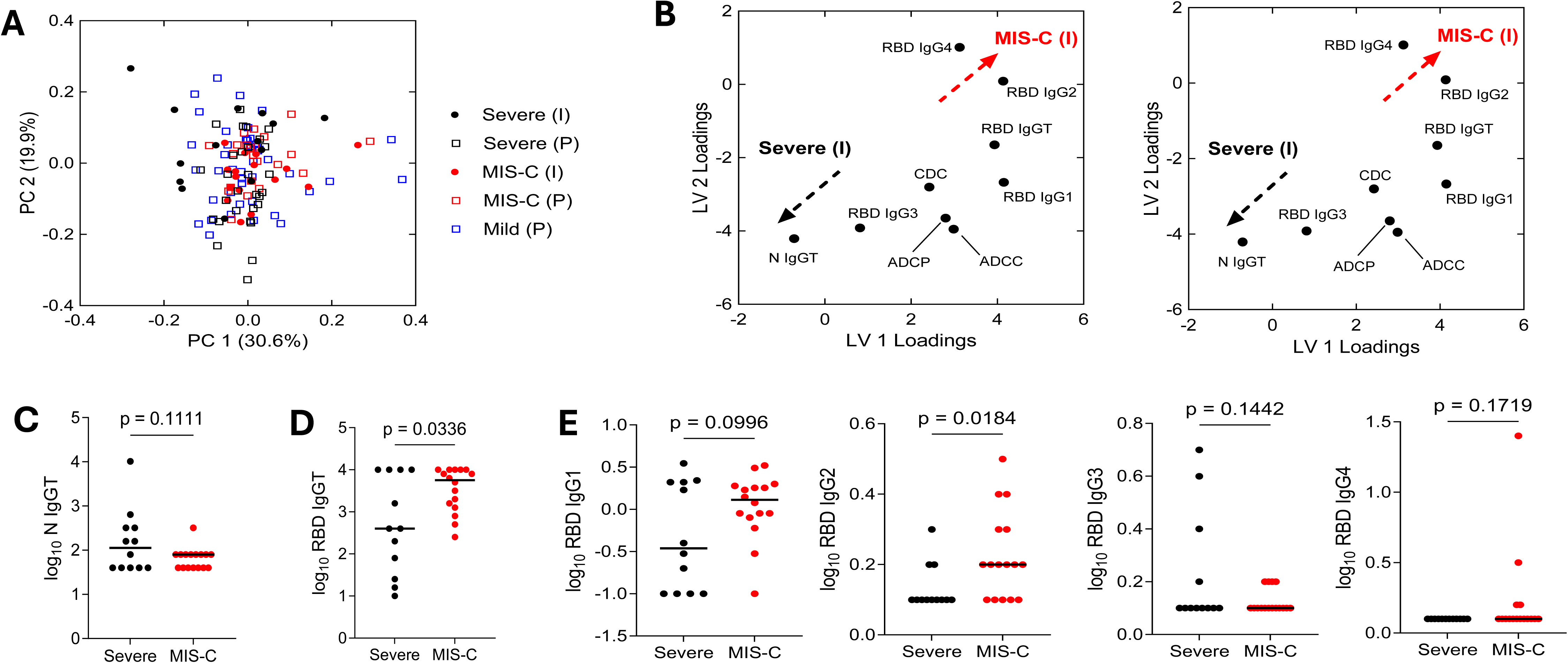
Anti-spike IgG antibody levels are higher, but Fc-mediated effector functions do not differ between incident MIS-C and Severe COVID-19 patients. For these analyses, a total of 107 samples were used (MIS-C, n = 39; Severe COVID-19, n = 26; Mild/asymptomatic infection, n = 42). (A) Principal component analysis of antibody levels and Fc-mediated effector functions from all patient cohorts. (B) Partial least squares discriminant analysis of antibody levels (OD_405_ values) and Fc-dependent effector functions (PLS-DA) scores (left) and loadings (right) for incident MIS-C and Severe COVID-19 patients. (C) Log_10_-transformed anti-Nucleocapsid total IgG OD_405_ values from incident MIS-C and Severe COVID-19 patients. (D) Log_10_-transformed anti-Spike RBD total IgG OD_405_ values for incident MIS-C and Severe COVID-19 patients. (E) Log_10_-transformed anti-Spike RBD IgG subclasses OD_405_ values for incident MIS-C and Severe COVID-19 patients. Black lines in (C), (D), and (E) denote median measurements for each group. Statistical tests were performed by Welch’s t-test. Abbreviations: I, incident; P, prevalent; RBD, spike receptor binding domain; ADCC, antibody-dependent cellular cytotoxicity; ADCP, antibody-dependent cellular phagocytosis; CDC, complement-dependent cytotoxicity; N, nucleocapsid; IgGT, total IgG.

Further analysis of IgG abundance showed no differences in anti-N antibody levels between the two groups (**Fig. 2C**). In contrast, anti-RBD IgG levels were higher in MIS-C than in severe COVID-19 (**Fig. 2D**). This difference was most apparent with IgG1 and IgG2 (**Fig. 2E**). The levels of IgG3 and IgG4 were at or below the detection limit of our assay (**Fig. 2E**). Fc-mediated effector functions remained indistinguishable between the two groups (**Supplementary Fig. S4**).

### Autoantibodies are more abundant in the SARS-CoV-2 infected patient groups compared to healthy controls

Autoantibodies have been shown to be induced in adults with severe COVID-19, and anticytokine antibodies specific for Type I interferons are pathogenic and associated with severe COVID-19 manifestations^1, 29^. We characterized autoantibody profiles in the three SARS-CoV-2 infection groups and healthy controls using a custom 73-plex autoantigen subarray (**Table S2).** Since 33 MIS-C patients had been administered IVIG, a treatment that contains pooled antibodies from healthy donors and obscures autoantibody analysis, we focused autoantibody analysis on MIS-C patients who were at least 100 days post-IVIG treatment (*n* = 21), when > 90% of exogenous immunoglobulin has been metabolized (**Fig. S2**). We found that children in all SARS-CoV-2 infection groups showed increased autoreactivity, regardless of clinical presentation, with ∼51% of infected subjects positive for at least 1 autoantibody, in contrast to only ∼14% of healthy controls (**Fig. 3A**, **Fig. S3**). Autoantibody levels were similar among mild/asymptomatic infection (55%), severe COVID-19 (50%), and MIS-C (39%) patients.

**Figure 3.**
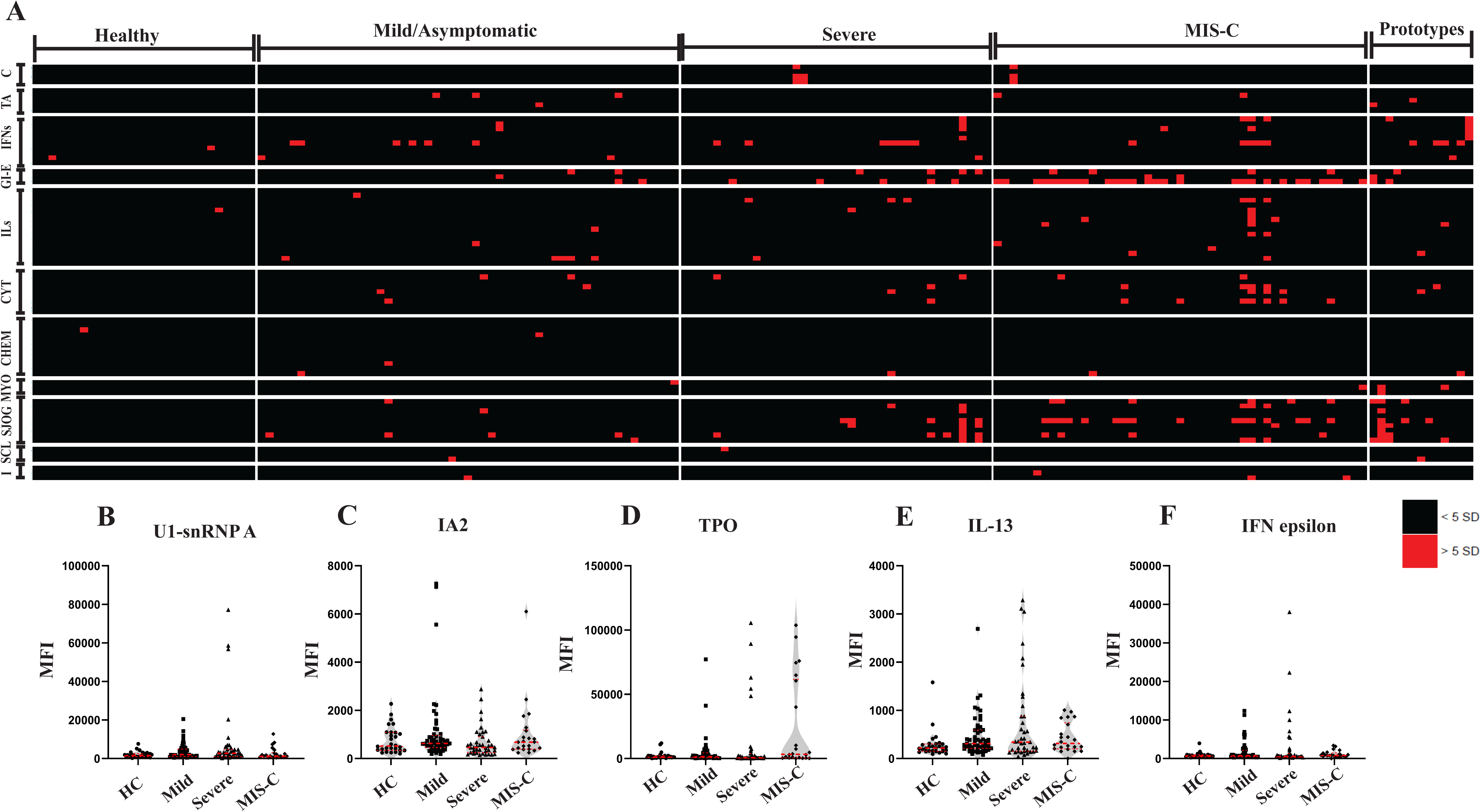
Higher prevalence of autoantibodies in SARS-CoV-2 infected subjects compared to healthy controls (HC). (A) Heatmap representing plasma IgG antibodies measured using a 73-plex, microbead-based protein array. Healthy controls (HC) (n = 28) and patients who were diagnosed with mild/asymptomatic SARS-CoV-2 infection (n = 53), severe COVID-19 (n = 39), MIS-C (n = 47), and prototypes (positive controls) (n = 13) are shown on the x-axis. Autoantigens are grouped on the y-axis based on disease: traditional autoantigens (TA), interferons (IFNs), gastrointestinal and endocrine disorders (GI-E), interleukins (ILs), other cytokines/growth factors/receptors (CYT), chemokines (CHEM); connective tissue diseases including myositis/overlap syndromes (MYO), and SLE/Sjögren’s syndrome (SJOG), and scleroderma (SCL), and antigens associated with tissue inflammation or stress responses (I). C, controls. Autoantigen names are provided in **Table S2**. Colors indicate autoantibodies whose MFI measurements are > 5 SD (red) or < 5 SD (black) above the average MFI for HC. MFIs <3000 were excluded. (B-C) Violin plots comparing MFI data for (B) autoantigens (U1-snRNPA; IA2; and Thyroperoxidase, TPO) and (C) prominent outlier cytokines (Interferon-epsilon and IL-13) among HC, mild SARS-CoV-2 infection, severe COVID-19, and MIS-C patients [only blood draws >100 days post IVIG treatment (n = 18)]. The dotted middle line represents the median, and the upper and lower dotted lines correspond to the first and third quartiles. MFI values are represented on the y-axis and MFI values for individual participants are displayed as dots for HC, squares for mild SARS-CoV-2 infection, triangles for severe COVID-19, and diamonds for MIS-C. Dunn-Bonferroni statistical analyses and corrections were performed to determine statistical significance between groups, with asterisks corresponding to the adjusted p values; **** p < 0.0001, *** p < 0.001,** p < 0.01, * p < 0.05. Non-significant comparisons (p > 0.05) are not shown. Source data are provided as a Source Data file.

COVID-19 has been associated with an increased risk of autoimmune disorders, especially connective tissue diseases^30^ ^31^. To explore this further, we analyzed connective tissue disease (CTD) associated autoantibodies commonly targeted in four distinct diseases (Scleroderma, Myositis/Overlap Syndrome, SLE, and Sjögren’s syndrome) across the SARS-CoV-2 infection groups and HC. We found that, collectively, 19% of SARS-CoV-2-infected patients – but none of the healthy control subjects -- were positive for at least one CTD autoantibody (**Fig. 3A**). CTD autoantibody prevalence was highest in patients with severe COVID-19 (25%), with similarly lower levels in mild/asymptomatic infection (15%) and MIS-C (∼17%). Moreover, approximately 15% of subjects across the infection groups tested positive for at least one autoantibody within the SLE/Sjögren’s CTD subpanel (11% of mild infection, 20% of severe COVID-19, and 17% of MIS-C). Fewer than 4% of infected patients in all groups tested positive for at least one autoantibody in the myositis and scleroderma subpanels. Five subjects (four in the severe COVID-19 group and one in the mild/asymptomatic infection group) were positive for multiple SLE/ Sjo gren’s associated self-antigens (**Table 2**). Indeed, two of these children had SLE. Notably, one tested positive for 13 autoantigens, seven of which were SLE/ Sjo gren’s subpanel autoantigens, suggesting profound autoimmune dysregulation in this individual.

**Table 2.** Clinical characteristics of patients with prominent autoantibodies.

| **Patient ID** | **Infection Status** | **Autoantibody** | **Age Range (Years)** | **Sex assigned at birth** | **Race** | **Vaccination status** | **Days between admission and collection** |
| --- | --- | --- | --- | --- | --- | --- | --- |
| MLD#18 | Mild | U1-snRNPA, La/SSB, TNF-alpha,CXCL8 | 12-17 | F | White | Y | 62 |
| SCD# 22 | Severe | Ro60/SSA, Sm/RNP | 6-11 | F | Black | N | 10 |
| SCD# 36 | Severe | IFN-alpha1, 2,8,10, Ribo P0, P1, Ro60/SSA, sm/RNP, Smith, U1-snRNP A, U1-snRNP C, Complement 3, PDC E2 | 6-11 | M | Hispanic/Latino | Y | 884 |
| SCD# 39 | Severe | IFN-lambda2, Ro60/SSA, Sm/RNP, U1-snRNP A, U1-snRNP C, PDC-E2 | 18-21 | F | Hispanic/Latino | Y | 305 |
| SCD# 27 | Severe | IFN-epsilon, IL-13, MIP-1-alpha, LIF, Ribo P0 | 12-17 | M | Hispanic/Latino | N | 4 |

When we analyzed particular autoantigens, we observed that U1-snRNP-A, a component of the U1-small nuclear ribonucleoprotein complex (U1-snRNP) and a target of autoreactive B cells and T cells in rheumatic diseases such as mixed connective tissue disease (MCTD) and systemic lupus erythematosus (SLE) ^32^, was a frequent outlier in patients with severe COVID-19 (**Fig. 3B**, left panel). Outliers were also identified exhibiting high IgG autoantibodies to IA2, an autoantibody typically associated with type 1 diabetes (T1D) ^33^, in the mild/asymptomatic infection and in MIS-C (Fig. 3B, middle panel), suggesting that these children may be at risk for glucose dysregulation or type 1 diabetes (**Fig. 3B**, middle panel). The GI/Endocrine subpanel was associated with a greater prevalence of autoantibodies in MIS-C patients relative to other subpanels and groups (**Fig. 3A**). High levels of autoantibodies in this subpanel were observed mostly in MIS-C patients (∼28%) and severe COVID-19 (∼23%), with fewer occurrences in the mild/asymptomatic infection group (∼8%). In particular, autoantibodies to thyroperoxidase (TPO), which is associated with autoimmune thyroid diseases, were common in MIS-C (22%) and severe COVID-19 (15%) and rare in the mild/asymptomatic infection group (∼4%) (**Fig. 3B**, right panel).

Severe SARS-CoV-2 disease has been associated with cytokine storm, a hyperinflammatory immune response characterized by massive release of cytokines ^34^. We explored whether high cytokine production was linked with increased production of autoantibodies to cytokines and found that approximately one-third (32%) of children in the SARS-CoV-2 infection groups exhibited autoreactivity to inflammatory cytokines, a proportion that is much higher than that observed in healthy controls (∼14%). Differences among infection groups were not significantly different, with anti-cytokine antibodies being higher in mild/asymptomatic infection (∼36%) than in severe COVID-19 (30%) and MIS-C (∼22%) (**Fig. 3A**). We also observed that anti-interleukin antibodies showed minimal variation across infection groups (ranging between 11% and 13% subjects with at least one autoantibody in each group). When we analyzed autoantibodies against IL-13, which has been reported as a driver of COVID-19 severity ^35^, overall levels of anti-IL-13 autoantibodies were higher in the mild/asymptomatic infection group than in healthy controls; however, high-antibody outliers were most common in the severe COVID-19 patients (**Fig. 3C**, left panel).

Interferons (IFN), especially Type 1 IFNs, play a crucial role in preventing viral replication of SARS-CoV-2 ^36^. Interferon autoreactivity was more commonly observed in children who had severe COVID-19 (∼23%) and mild/asymptomatic infection (17%) compared to healthy children (∼7%). No children diagnosed with MIS-C were positive for interferon autoantibodies. In particular, IFN-epsilon, an essential antigen involved in anti-viral activity ^37^, was found to be a target of autoantibodies in severe COVID-19 (18%) and mild/asymptomatic infection (11%) but not in the MIS-C and HC groups (**Fig. 3C**, right panel). Except for two subjects with severe COVID-19 (SCD #22 and SCD#36) exhibiting an autoimmune response to multiple interferons (**Table 2**), subjects who tested positive for interferon autoantibodies displayed reactivity to only one interferon.

## Discussion

We report that the clinical spectrum of SARS-CoV-2 infection in children is associated with a heterogeneous repertoire of antibodies against SARS-CoV-2 antigens and autoantibodies. Among anti-SARS-CoV-2 antibodies, our data are consistent with links between antibody levels to immunodominant viral antigens, antigen load, immune protection, and clinical severity. Our results also show that levels of IgG antibodies against the receptor binding domain of the viral spike protein do not correspond to non-neutralizing antibody effector functions, suggesting yet another aspect of virus-induced immune dysfunction. Among antibodies to autoantigens, we show SARS-CoV-2-induced autoreactivity, particularly against connective tissue disease antigens, some cytokines, and the type I interferon, IFN-ε. We also show nuanced differences in the autoreactivity across clinical presentations of SARS-CoV-2 infection, with some autoantibodies emerging as markers of clinical severity. Together, our results contribute to elucidating the antibody repertoire generated in response to pediatric SARS-CoV-2 infection, its association with clinical disease severity, and to hypotheses on causative relationships between clinical presentation of SARS-CoV-2 infection and autoreactivity.

Multiple considerations derive from the analysis of antibodies directed to the immunodominant SARS-CoV-2 antigens, Spike (S) and Nucleocapsid (N). First, when we analyzed data by infection group, regardless of time interval between infection diagnosis and time of sample collection, we found that anti-N antibodies tended to be higher in severe COVID-19 than in MIS-C. This result was observed with both assay platforms in the study (multiplex bead array and ELISA) and different approaches to data analysis. Since the immunodominant N protein is among the most abundant structural proteins in SARS-CoV-2-infected cells,^38, 39^ these data suggest an association between anti N antibody production and viral load, which can be expected to be higher in severe COVID-19 than in MIS-C, since the latter is a post-acute manifestation. Previous reports that anti-N IgG antibody levels correlate with COVID-19 disease severity ^40^ are consistent with our interpretation. Second, higher anti-S antibody levels in subjects with mild/asymptomatic infection than those with severe COVID-19 may correlate with immune protection, since it is well established that the neutralizing antibodies primarily target SARS-CoV-2 Spike and its RBD, which directly interacts with the ACE2 receptor on host cells^41, 42, 43^. Further analysis of anti-RBD antibodies also showed that, at least in samples collected during or shortly after hospitalization, anti-S antibody levels were significantly higher in the MIS-C than in the severe COVID-19 group. This result agrees with some previous observations ^44^ and disagrees with others that, for example, found increased anti-RBD levels in MIS-C children only in the IgA fraction ^45^. These differences may relate to differences in sample sizes and study design, particularly those concerning COVID-19 vaccination status and the clinical characteristics of the comparator groups. Third, in contrast with differences in IgG abundance between incident MIS-C and severe COVID-19 groups, which can be ascribed mostly to IgG1, we found no differences in Fc-mediated effector functions between these two groups. This is surprising, since IgG1, which is the most abundant IgG subclass, can be associated with most of the Fc-mediated effector functions investigated in our study^46^. The other subclass that exhibits such functions, IgG3, was barely detectable in most samples. Thus, the observed discrepancy between relative abundance and expression of function suggests that MIS-C may be associated with abnormal antibody function, due to, for example, altered IgG glycosylation patterns ^11, 12^. Additional studies are required to assess the links between antibody structure and function, hyperinflammation, and MIS-C outcome.

Virus-driven autoimmunity has been widely reported across various viral pathogens and arises through multiple mechanisms, including epitope spreading, molecular mimicry, and bystander activation ^47, 48^. SARS-CoV-2 has been implicated in triggering new autoantibodies in severely ill patients^1^ and classifiable autoimmune disease in previously healthy individuals ^31, 49, 50^, while patients with preexisting autoantibodies against type I interferons may be more susceptible to a severe disease outcome ^29^. The dual role of autoimmunity—both as a consequence of SARS-CoV-2 infection and as a factor worsening disease severity—underscores its multifaceted nature in SARS-CoV-2-induced pathology^29^.

We utilized a custom autoantigen array to examine the autoantibody repertoire of children with varying clinical manifestations of SARS-CoV-2 infection to further explore the role of autoimmunity in COVID-19. Our data indicates a link between CTDs and SARS-CoV-2, evidenced by the detection of CTD-specific autoantibodies in infected patients, regardless of the degree of clinical severity, and associated with lack of reactivity in healthy controls. This finding aligns with current literature that has implicated SARS-CoV-2 as a risk factor for autoinflammatory connective tissue disorders^16^. Nonetheless, without longitudinal samples, including those prior to infection, it remains uncertain whether the observed autoreactivity results from the SARS-CoV-2 infection or rather from preclinical autoimmunity culminating in severe disease outcome. In contrast to CTD autoreactivity, cytokine autoreactivity was observed in the healthy controls, albeit at half the frequency seen in the infected children. This can be expected as healthy individuals possess a unique autoreactome or “natural autoantibodies” -- unrelated to recent viral infections or underlying autoimmune disease -- that contribute to immune regulation ^51^. The increase in cytokine autoreactivity in infected individuals could be attributed to SARS-CoV-2-induced cytokine storm, an excessive immune response characterized by the unmitigated release of cytokines ^19^. Studies of longitudinal samples, including pre-SARS-CoV-2-infection sampling, are needed to test this possibility.

While autoimmune responses differed between infected and healthy (uninfected) children, differences among the various clinical presentations of SARS-CoV-2 infection were more nuanced. In general, the presence of autoantibodies directed against cytokines was similar across infection groups. However, certain autoantibodies emerged as distinct markers associated with specific clinical manifestations. For example, anti-IA2 antibodies were present in several patients with a mild presentation of COVID-19, a finding not observed in those with severe infections and only rarely seen in MIS-C. Moreover, children who developed MIS-C were more likely than any other infection group to produce autoantibodies against TPO. Yet, anti-TPO antibodies were also frequent in the severe COVID-19 group. These results suggest that these autoantibodies are a shared feature of immune dysregulation across both severe infection and post-infection states. Specific autoantibodies associated with severe COVID-19 included IL-13, which has been identified as a driver of disease ^21^.

Furthermore, U1-snRNP-A emerged as a prominent antigen in both mild and severe infections; however, it displayed heightened reactivity in multiple patients with severe disease. This suggests that U1-snRNP-A autoantibodies might serve as a marker of immune dysregulation rather than a distinct driver of disease. These data collectively suggest the possibility that COVID-19 is a risk factor for long-term autoimmune complications by acting as both a trigger and amplifier of autoimmunity.

Our study has limitations. The sample size in the study limits our ability to stratify results in each infection group by degree of clinical severity and age of study participants. This is important because, for example, MIS-C may present with relatively mild to very severe clinical symptoms ^12, 52^, which may be associated with different degrees of immune dysfunction, as we have proposed elsewhere^53^. Similar sample size considerations also apply to age groups, since the immune system evolves with age^54^. Additionally, the cross-sectional study design is not appropriate to investigate the duration and temporal evolution of antibody responses in the conditions under study. Moreover, the absence of pre-infection samples limits our ability to ascertain whether the autoantibodies detected in some study participants resulted from SARS-CoV-2-induced immune dysregulation or were associated with preexisting autoimmune conditions. Lastly, since blood was collected after IVIG treatment of MIS-C, analysis of autoantibodies in this patient group was limited to subjects who were at least 100 days post-IVIG treatment, when most exogenous immunoglobulin has been metabolized, thus reducing sample size. Despite these limitations, however, the data are robust and suggest correlations between clinical presentations and antibody responses that warrant further investigations.

In conclusion, this study shows that profiles of antibodies against SARS-CoV-2 and autoantigens vary with the clinical presentation of SARS-CoV-2 in children. In particular, our data suggest that antibody responses differ between severe COVID-19 and MIS-C. Further research and large studies are needed to unravel the complexities of the antibody responses to SARS-CoV-2 infection in children and to identify potential targets for therapeutic intervention and disease management, which may have implications for the understanding of Long COVID pathogenesis, prevention, and treatment.

## Data Availability

All data produced in the present study are available upon reasonable request to the authors

## Acknowledgments.

We thank Charles Hevi for coordinating biospecimen collection and shipping to biorepository with consortium members; and Beth Dworetzky and Steph Lomangino from Family Voices for their assistance and guidance with recruitment materials, surveys, and other participant-facing documents. We are indebted to patients and their families for consenting to participate to the study. This research was funded in part by NICHD R68/R33HD105619 and R33HD105593-03S2, NIAID R01AI158911, NCATS UL1TR003017, NHLBI R38HL143615, and NIH Agreement OT2 HL161847 through the NIH RECOVER Pathobiology Research Program. PJU was also supported by the Henry Gustav Floren Trust; Stanford Department of Medicine Team Science Program; Stanford Medicine Office of the Dean; NIAID R01AI175771 and R01AI182319-02. The views and conclusions contained in this document are those of the authors and should not be interpreted as representing the official policies, either expressed or implied, of the NIH.

