## Supplementary Information for "Antibody repertoire associated with clinically diverse presentations of pediatric SARS-CoV-2 infection"

**Supplementary Figures**

**Figure S1. Plasma IgG antibodies against viral proteins vary among SARS-CoV-2 infection severities.** (A) Heatmap representing plasma IgG measured using a 42-plex protein array. Healthy controls (HC) (n = 28), and participants who were diagnosed with Mild/Asymptomatic SARS-CoV-2 infection (n = 53), Severe COVID-19 (n = 39), MIS-C (n = 47), and prototypes (positive controls) (n = 5) are shown on the x-axis. Antigens are grouped on the y-axis by Control antigens and SARS-CoV-2 subgroups: non-structural proteins (NSP); accessory proteins (AP), and structural proteins; Other viral antigens, including EBV. Colors indicate IgG antibodies whose MFI measurements are > 5 standard deviation (SD) (red) or < 5 SD (black) above the average MFI for HC. MFI values <3000 were excluded. (B-E) Violin plots showing MFI data in the diagnostic groups in panel A. In these panels, MIS-C patients were only those at least 100 days post IVIG treatment (n = 18). The dotted middle line is the median, and the upper and lower dotted lines correspond to the ﬁrst and third quartiles. MFI values are represented on the y-axis. MFI values for individual participants are displayed as dots for HC, squares for mild infection, triangles for severe COVID-19, and diamonds for MIS-C. The violin plots show IgG levels against viral proteins that (B) are higher in all SARS-CoV-2 diagnostic groups relative to HCs; (C) are lower in all SARS-CoV-2 diagnostic groups relative to HCs; (D) show no differences across groups. (E) shows levels of IgG antibodies against nonstructural SARS-CoV-2 proteins. Dunn-Bonferroni statistical analyses and corrections were performed to determine statistical signiﬁcance between cohorts, with asterisks corresponding to the adjusted p values; **** p <0.0001, *** p <0.001, ** p <0.01, * p <0.05. Non-signiﬁcant comparisons (p > 0.05) are not shown. Source data are provided as a Source Data ﬁle.

**Figure S2. Presence of autoantibodies in patients with MIS-C is affected by time since IVIG administration.** Healthy controls and MIS-C patients grouped by time between hospital admission and blood collection [ < 1 week (n = 14), 1 week – 2 months (n = 12), > 2 months (n = 21)] and prototypes (positive controls) (n = 13) are shown on the x-axis. Autoantigens are grouped on the y-axis based on disease: traditional autoantigens (TA), interferons (IFNs), gastrointestinal and endocrine disorders (GI-E), interleukins (ILs), other cytokines/growth factors/receptors (CYT), chemokines (CHEM); connective tissue diseases including myositis/overlap syndromes (MYO), and SLE/Sjögren’s syndrome (SJOG), and scleroderma (SCL), and antigens associated with tissue inﬂammation or stress responses (I). C, controls. Colors indicate autoantibodies whose MFI measurements are > 5 SD (red) or < 5 SD (black) above the average MFI for HC. MFIs <3000 were excluded. Source data are provided as a Source Data ﬁle corresponding to Fig. 2.

**Figure S3. Higher prevalence of autoantibodies in patients infected with SARS-CoV-2 compared to healthy controls (HC).** Heatmap representing plasma IgG autoantibodies measured using a 77-plex, microbead-based protein array. Healthy controls (n = 28), and participants who were diagnosed with Mild/Asymptomatic SARS-CoV-2 infection (n = 53), Severe COVID-19 (n = 39), MIS-C (n = 47), and prototypes (positive controls) (n = 13) are shown on the x-axis. Autoantigens are grouped on the y-axis as in Fig. S2. Colors correspond to the MFI gradient values. Source data are provided as a Source Data ﬁle corresponding to Figure 2.

**Figure S4. Fc-mediated effector functions did not differ between incident MIS-C and severe COVID-19 patients.** Fc-mediated effector functions were examined for incident MIS-C (n=12) and severe COVID-19 (n=16) subjects: (A) antibody-dependent cellular cytotoxicity (ADCC); (B) antibody-dependent cellular phagocytosis (ADCP); (C) Complement-dependent cytotoxicity (CDC). Black lines in (A), (B), and (C) denote median measurements per group. Statistical tests were performed by Welch’s t-test. ns, non significant.

**Supplementary Tables**

**Supplementary Table 1.** Viral antigen array content.

**Supplementary Table 2.** Autoantigen array content
