## Supplementary figures and images for "Antibody repertoire associated with clinically diverse presentations of pediatric SARS-CoV-2 infection"

### Supplementary Figures

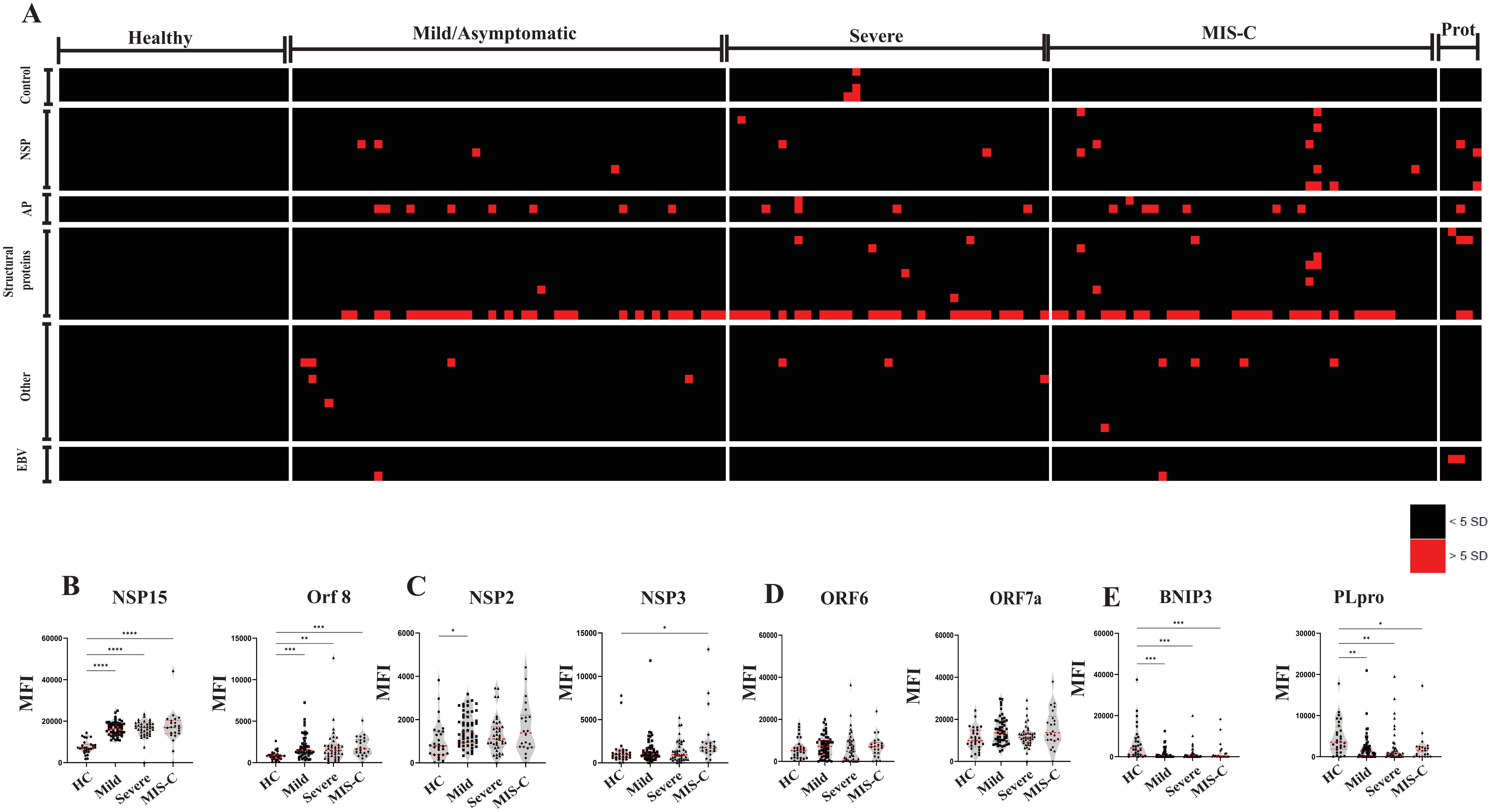

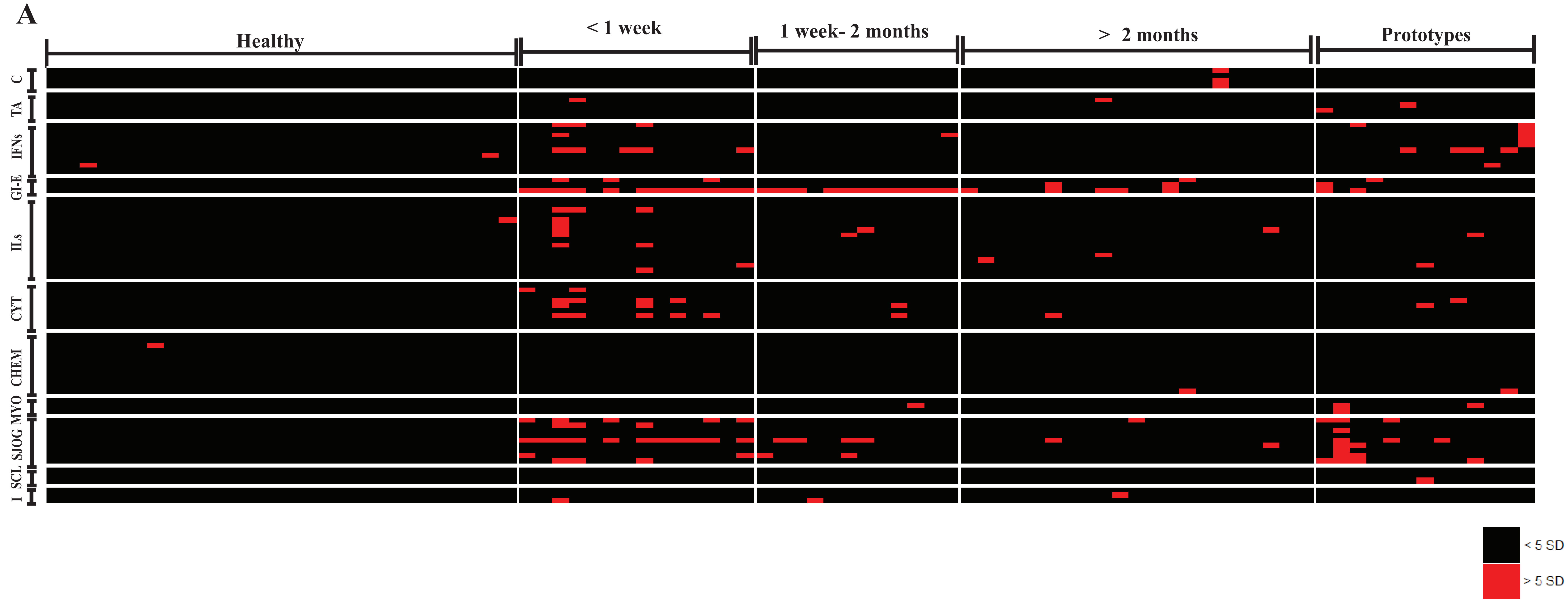

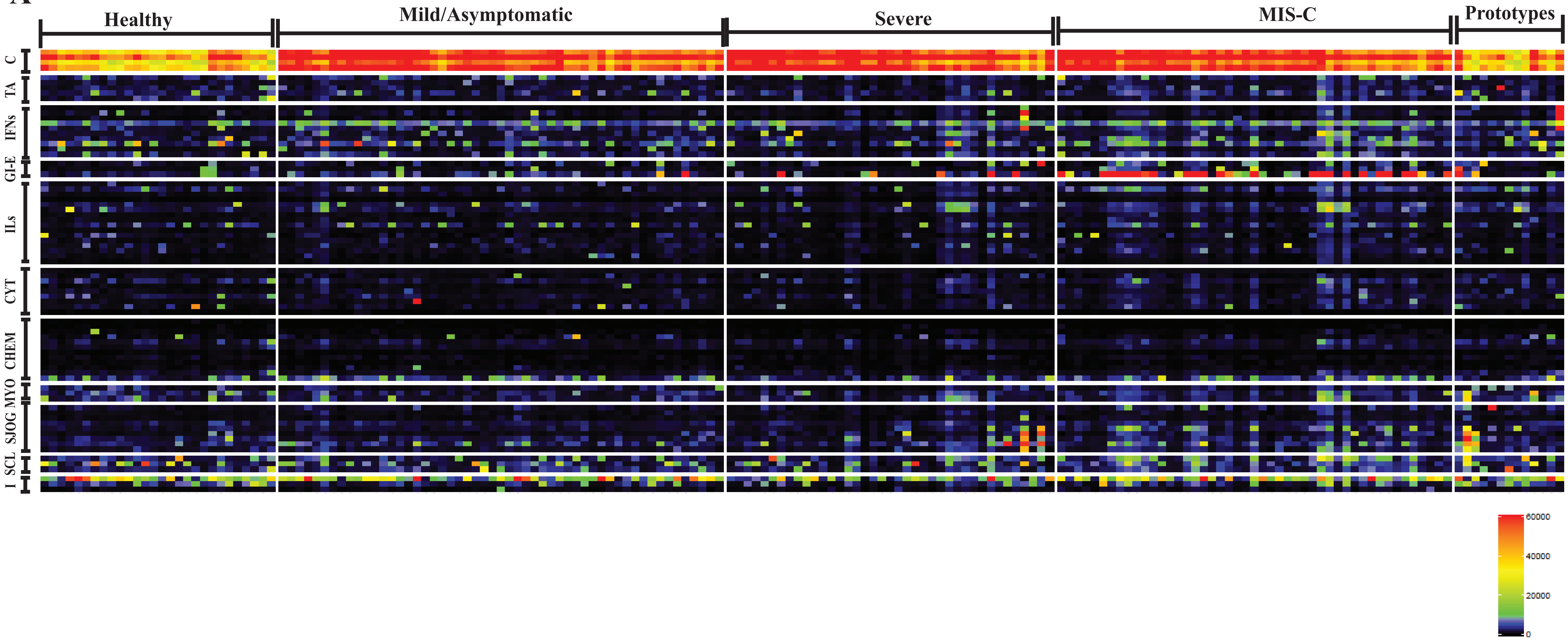

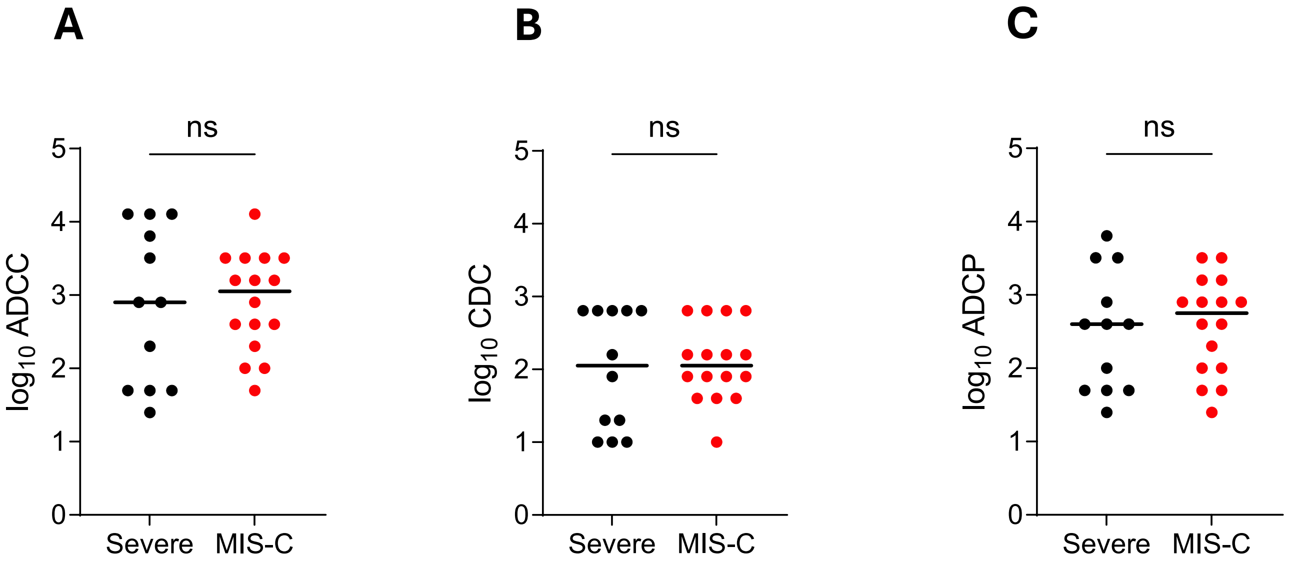
