## Supplementary material for "Antibody repertoire associated with clinically diverse presentations of pediatric SARS-CoV-2 infection": Table S1

**Supplementary Table 1.** Viral antigen array content.

| **Bead ID** | **Antigen** | **Category** | **Vendor** | **Catalog #** |
| --- | --- | --- | --- | --- |
| 1 | Bare Bead |  |  |  |
| 2 | Anti-Human IgG Fc fragment specific | Control | Jackson | 109-005-008 |
| 3 | Anti-Human IgG F(ab’) fragment specific | Control | Jackson | 109-005-006 |
| 7 | Anti-Human IgG (H+L) | Control | Jackson | 109-005-003 |
| 8 | Human IgG from Serum | Control | Jackson | I4506 |
| 63 | ORF6 | Accessory Protein (AP) | ProSci | 10-437 |
| 99 | HMPV Glycoprotein G | Other | Sino | 40791-V08H |
| 100 | Influenza A hemagglutinin (HA) | Other |  |  |
| 112 | RSV-F | Other | Sino | 11049-V08B |
| 114 | EBV p18 | EBV | Prospec | Ebv-277 |
| 122 | EBV EA-D | EBV | MyBioSource | MBS319448 |
| 125 | Parainfluenza Hemagglutinin-neuraminidase | Other | Sino | 40629-V07B |
| 127 | CMV gB | Other | Prospec | CMV-211 |
| 129 | SARS2 S1 | Structural Protein | ProSci | 97-087 |
| 133 | SARS2 Spike RBD | Structural Protein | Sino | 40592-V08B |
| 142 | Rhinovirus VP1 | Structural Protein | MyBioSource | MBS1220686 |
| 143 | Spike S2 ECD | Structural Protein | ProSci | 10-115 |
| 145 | Nucleocapsid | Structural Protein | ProSci | 10-434 |
| 146 | Envelope | Structural Protein | ProSci | 97-082 |
| 149 | Membrane | Structural Protein | ProSci | 10-429 |
| 152 | NSP3 | Non-structural proteins (NSP) | ProSci | 10-406 |
| 153 | ORF7a | Accessory Protein (AP) | ProSci | 10-435 |
| 154 | ORF8 | Accessory Protein (AP) | ProSci | 10-002 |
| 156 | AcmNPV-gp64 Protein | Other | Sino | 40496-V08B |
| 157 | BNIP3 | Other |  |  |
| 158 | NSP2 | Non-structural proteins (NSP) | ProSci | 10-425 |
| 159 | Measles Nucleoprotein | Other | Prospec | MMP-001 |
| 161 | 3CLpro | Structural Protein | Sino | 40594-V56E |
| 163 | NSP8 | Non-structural proteins (NSP) | ProSci | 10-145 |
| 169 | NSP9 | Non-structural proteins (NSP) | ProSci | 10-417 |
| 170 | NSP10 | Non-structural proteins (NSP) | ProSci | 10-408 |
| 174 | NSP12 | Non-structural proteins (NSP) | R&D | 10686-CV |
| 176 | NSP13 | Non-structural proteins (NSP) | ProSci | 10-427 |
| 181 | NSP15 | Non-structural proteins (NSP) | ProSci | 20-207 |
| 182 | NSP16 | Non-structural proteins (NSP) | MyBioSource | MBS156017 |
| 186 | Methyltransferase | Structural Protein | Sino | 40598-V07E |
| 189 | SARS2 Helicase | Structural Protein | Sino | 40596-V07E |
| 191 | SARS2 RDRP | Structural Protein | Sino | 40595-V08B |
| 192 | NSP7 | Non-structural proteins (NSP) | Origene | TP750200 |
| 197 | Mumps | Other | Prospec | MMP-001 |
| 199 | Rubella | Other | Prospec | RUB-291 |
| 204 | EBV EBNA-1 | EBV | Abcam | Ab138345 |
| 216 | Plpro | Structural Protein | Sino | 40593-V07E |
| 284 | EV VLP1 (Enterovirus) | Other | BioMart |  |
| 285 | HBSAg | Other | MyBioSource | MBS142509 |
| 309 | Varicella Zoster Glycoprotein E | Other | MyBioSource | MBS553197 |
