## Supplementary material for "Antibody repertoire associated with clinically diverse presentations of pediatric SARS-CoV-2 infection": Table S2

**Supplementary Table 2.** Autoantigen array content.

| **Bead ID** | **Antigen** | **Category** | **Vendor** | **Catalog #** |
| --- | --- | --- | --- | --- |
| 1 | Bare Bead |  |  |  |
| 2 | Anti-Human IgG Fc fragment specific | Control (C) | Jackson | 109-005-008 |
| 4 | Anti-Human IgG F(ab’) fragment specific | Control (C) | Jackson | 109-005-006 |
| 6 | IFN-alpha1 | Control (C) | Prospec | CYT-291 |
| 7 | Anti-Human IgG (H+L) | Control (C) | Jackson | 109-005-003 |
| 8 | Human IgG from Serum | Control (C) | Jackson | I4506 |
| 12 | Complement 3 | Other Cytokine (CYTK) | Complement tech | A113 |
| 13 | CXCL10 | Chemokine (CHEM) | Peprotech | 300-12 |
| 15 | CCL26 | Chemokine (CHEM) | Peprotech | 300-48 |
| 18 | IFN-alpha10 | Interferon (IFNs) | Sino | 10349-H08H |
| 19 | IFN-epsilon | Interferon (IFNs) | R&D | 9667-ME-025/CF |
| 20 | CXCL9 | Chemokine (CHEM) | Peprotech | 300-26 |
| 25 | FGF7 | Other Cytokine (CYTK) | Peprotech | 100-19 |
| 26 | IL-10 | Interleukins (ILs) | Peprotech | 200-10 |
| 28 | GM-CSF | Other Cytokine (CYTK) | Peprotech | 300-03 |
| 29 | ACE2 | Other Cytokine (CYTK) | Sino | 10108-H05H |
| 30 | IL-13 | Interleukins (ILs) | Peprotech | 200-13 |
| 33 | IFN-alpha2 | Interferon (IFNs) | R&D | 11101-2 |
| 34 | IL-23 | Interleukins (ILs) | Peprotech | 200-01A |
| 35 | IL-1RA | Interleukins (ILs) | Peprotech | 200-01RA |
| 38 | IFN-alpha6 | Interferon (IFNs) | Origene | TP760329 |
| 39 | IFN-gamma | Interferon (IFNs) | Peprotech | 300-02 |
| 44 | IL-23 | Interleukins (ILs) | Peprotech | 200-23 |
| 45 | IL-11 | Interleukins (ILs) | Peprotech | 200-11 |
| 46 | IL-17A | Interleukins (ILs) | Peprotech | 200-17 |
| 47 | IL-17F | Interleukins (ILs) | Peprotech | 200-25 |
| 48 | IL-21 | Interleukins (ILs) | Peprotech | 200-21 |
| 49 | IL-22 | Interleukins (ILs) | Peprotech | 200-22 |
| 51 | IL-31 | Interleukins (ILs) | Prospec | CYT-625 |
| 53 | IL-4 | Interleukins (ILs) | Peprotech | 200-04 |
| 55 | IL-6 | Interleukins (ILs) | Peprotech | 200-06 |
| 55 | IL-7 | Interleukins (ILs) | Prospec | CYT-214 |
| 56 | IL33 | Interleukins (ILs) | Peprotech | 200-33 |
| 57 | IFN-lambda1 | Interferon (IFNs) | Peprotech | 300-02L |
| 58 | LIF | Other Cytokine (CYTK) | Peprotech | 300-05 |
| 59 | IFN-omega | Interferon (IFNs) | Peprotech | 300-02J |
| 60 | MIP-1alpha | Chemokine (CHEM) | Peprotech | 300-08 |
| 61 | PDGFBB | Other Cytokine (CYTK) | Peprotech | 100-14B |
| 63 | s-rank ligand | Other Cytokine (CYTK) | Peprotech | 310-01C |
| 65 | VEGFA | Other Cytokine (CYTK) | Peprotech | 100-20A |
| 66 | BPI | Inflammation (I) | Sigma | SRP6307 |
| 67 | TNF-alpha | Other Cytokine (CYTK) | Peprotech | 300-01A |
| 68 | C1q | Inflammation (I) | EMD | 204876 |
| 69 | PMScl-75 | Myositis/Overlap Syndromes (MYO) | Surmodics | A17001 |
| 70 | CENPA | Scleroderma (SCL) | Surmodics | A16901 |
| 72 | FBL | Scleroderma (SCL) | Prospec | ENZ-566 |
| 99 | PDC-E2 | Myositis/Overlap Syndromes (MYO) | Surmodics | A17901 |
| 100 | EJ | Myositis/Overlap Syndromes (MYO) | Surmodics | A11101 |
| 110 | IFN-lambda2 | Interferon (IFNs) | Peprotech | 300-02K |
| 112 | U1-snRNP C | SLE/ Sjogrens (SJOG) | Surmodics | A13201 |
| 114 | CXCL13 | Chemokine (CHEM) | Peprotech | 300-47 |
| 122 | CXCL16 | Chemokine (CHEM) | Peprotech | 300-55 |
| 125 | TPO | GI Endocrine (GI-E) | Surmodics | A12101 |
| 127 | CXCL5 | Chemokine (CHEM) | Peprotech | 300-22 |
| 129 | CCL21 | Chemokine (CHEM) | Peprotech | 300-35A |
| 133 | CXCL8 | Chemokine (CHEM) | Peprotech | 200-08 |
| 142 | IFN-alpha8 | Interferon (IFNs) | Sino | 10347-H08H |
| 143 | CCL22 | Chemokine (CHEM) | Peprotech | 300-36 |
| 145 | CCL19 | Chemokine (CHEM) | Peprotech | 300-29B |
| 146 | CCL25 | Chemokine (CHEM) | Peprotech | 300-45 |
| 156 | Jo-1 | Traditional Autoantigens (TA) | Surmodics | A12901 |
| 157 | Histone 1 | Traditional Autoantigens (TA) | Immunovision | HIS-1001 |
| 159 | Ribo P1 | SLE/ Sjogrens (SJOG) | Surmodics | A14201 |
| 197 | IL-15 | Interleukins (ILs) | Peprotech | 200-15 |
| 199 | Ribo P2 | SLE/ Sjogrens (SJOG) | Surmodics | A14301 |
| 235 | Smith | SLE/ Sjogrens (SJOG) | Immunovision | SMA-3000 |
| 284 | IA2 | Traditional Autoantigens (TA) | Novus |  |
